# Interplay of Type 2 Inflammation, Small Airway Dysfunction, and Patient-Reported Outcomes in Treatment-Naïve Uncontrolled Asthma: A Retrospective Primary Care Analysis

**DOI:** 10.1101/2025.02.18.25322437

**Authors:** Yutaka Nakano, Rika Nakano, Takuya Yukawa, Akihiro Arita

**Affiliations:** Nakano Respiratory and Allergy Clinic, 381-2 Hatsuoi-cho, Chuo Ward, Hamamatsu, Shizuoka, 433-8112 Japan; AOI Pharmacy, Hatsuoiten, 381-8 Hatsuoi-cho, Chuo Ward, Hamamatsu, Shizuoka, 433-8112 Japan

**Keywords:** asthma, biomarkers, eosinophils, nitric oxide, patient-reported outcome measures, small airway dysfunction, spirometry

## Abstract

**Background:** Effective initial asthma treatment in primary care requires a comprehensive assessment of airway inflammation and lung function impairment, including small airway dysfunction (SAD). We investigated the prevalence of type 2 (T2) inflammation and SAD in treatment-naïve patients with uncontrolled asthma who received primary care, and assessed the utility of the Asthma Control Questionnaire-5 (ACQ-5) as a potential indicator of these factors.

**Methods:** This single-center retrospective study enrolled treatment-naïve adults who presented with uncontrolled asthma (ACQ-5 score ≥ 1.5) between April 2020 and March 2022. T2 inflammation was assessed using blood eosinophil count (bEOS), fractional exhaled nitric oxide (FeNO), and total immunoglobulin E (IgE) levels. Patients with bEOS ≥ 300 cells/μL and FeNO ≥ 50 ppb were designated “T2-high.” SAD was defined as a forced expiratory flow between 25% and 75% of the vital capacity percentage predicted value (FEF_25-75_%pred) < 65%.

**Results:** Among 192 patients, 87% exhibited an elevation in at least one T2 biomarker, with 47% meeting the T2-high criteria. An ACQ-5 score ≥ 3.0 significantly predicted T2-high status (odds ratio: 2.62, 95% confidence interval: 1.46-4.69, p = 0.00127). SAD was identified in 52% of patients, with ACQ-5 scores showing a strong negative correlation with FEF_25-75_%pred (ρ = -0.583, p < 0.0001), particularly for nocturnal awakening score (ρ = -0.728, p < 0.0001).

**Conclusions:** Our findings revealed a high prevalence of T2 inflammation and SAD in treatment-naïve patients with uncontrolled asthma who received primary care. The ACQ-5 demonstrates potential as a practical screening tool for these pathophysiological features, offering valuable guidance for initial treatment decisions where advanced diagnostic capabilities may be limited.

**Data availability statement:** Data are available upon reasonable request. The datasets used and/or analyzed during the current study are available from the corresponding author upon reasonable request. http://creativecommons.org/licenses/by-nc/4.0/

This is an open-access article distributed in accordance with the Creative Commons Attribution Noncommercial (CC BY-NC 4.0) license, which permits others to distribute, remix, adapt, build upon this work noncommercially, and license their derivative works on different terms, provided that the original work is properly cited, appropriate credit is given, any changes are indicated, and the use is noncommercial.

See: http://creativecommons.org/licenses/by-nc/4.0/. https://doi.org/10.5061/dryad.j6q573np9

**What is already known on this topic:** Despite therapeutic advances, optimal asthma control remains elusive in many adult patients, particularly in primary care settings, where most treatments occur. Our understanding of type 2 inflammation and small airway dysfunction in treatment-naïve patients with uncontrolled asthma remains limited. This knowledge gap hinders the development of enhanced management strategies among primary care practitioners.

**What this study adds:** This study revealed high prevalence rates of type 2 inflammation (87%) and small airway dysfunction (52%) in treatment-naïve patients with uncontrolled asthma receiving primary care, with 47% showing high-grade type 2 inflammation. An Asthma Control Questionnaire-5 (ACQ-5) score ≥ 3.0, predicted high-grade type 2 inflammation, while nocturnal symptoms correlated with small airway dysfunction.

**How this study might affect research, practice, or policy:** These findings underscore the importance of considering type 2 inflammation and small airway dysfunction when initiating asthma treatment in primary care. ACQ-5 could serve as a practical tool for more targeted treatment decisions, while future research should focus on developing primary care-specific algorithms that combine ACQ-5 scores with biomarker profiles.

## Introduction

Asthma affects over 350 million people worldwide and imposes a significant burden on healthcare systems due to frequent exacerbations and hospitalizations.^1^ Despite substantial advancements in understanding its pathophysiology and the widespread adoption of evidence-based guidelines, approximately 40% of adults with asthma remain inadequately controlled,^2–4^ highlighting the need for improved management strategies in primary care settings, where most patients receive their initial treatment.^5^ Building on recent findings, this study aimed to clarify the prevalence of type 2 (T2) inflammation^6^ and small airway dysfunction (SAD)^7^ in a primary care setting and assess the utility of the Asthma Control Questionnaire-5 (ACQ-5)^8^ scores as indicators of these conditions, an area not comprehensively explored in treatment-naïve patients.

Recent evidence supports a personalized medical approach that targets “treatable traits”^9,10^ rather than employing a one-size-fits-all treatment strategy.^11^ Among these traits, two key pathophysiological mechanisms, T2 inflammation and SAD, play crucial roles in the initial evaluation in primary care settings.^12^

T2 inflammation, driven by cytokines such as interleukin (IL)-4, IL-5, and IL-13, is central to the pathogenesis.^13^ Biomarkers of T2 inflammation not only predict the risk of exacerbations but also guide the use of biological therapies in severe asthma.^14–19^ Notably, recent studies suggest that high-dose inhaled corticosteroids (ICS) provide greater benefits for patients with elevated T2 biomarkers who have previously been treated but remain uncontrolled compared with medium-dose therapy.^20, 21^ This highlights the potential value of biomarker-guided treatment selection in the initial treatment of treatment-naïve patients with uncontrolled asthma.

Similarly, SAD is increasingly being recognized as a contributor to poor asthma control, frequent exacerbations, and impaired quality of life.^22, 23^ Despite its clinical significance, the prevalence and impact of SAD in treatment-naïve patients remain poorly characterized in primary care, largely because of the limited implementation and availability of spirometry and other diagnostic tools.

The ACQ-5 is a widely validated tool in clinical practice that assesses asthma control based on five key symptom-related items, including frequency of symptoms, nocturnal awakenings, and activity limitation.^8^ The ACQ has demonstrated reliability and responsiveness, making it suitable not only for clinical management but also for research and quality of care assessments in primary care settings.^24^ While primarily designed to evaluate symptom control, its potential utility as an indirect marker of underlying pathophysiological mechanisms, such as T2 inflammation and SAD, remains insufficiently investigated, particularly in treatment-naïve patients within primary care, where access to advanced diagnostic modalities is often limited.

Given these challenges and the urgent need for practical and accessible tools to guide personalized asthma treatment in primary care, this study aimed to address these questions.

1. What is the prevalence of elevated T2 biomarkers and SAD in treatment-naïve patients with uncontrolled asthma receiving primary care?
2. What proportion of these patients may benefit from the initial high-dose ICS therapy based on their T2 inflammatory status?
3. How are T2 biomarkers, ACQ-5 scores, and lung function parameters, particularly SAD, interrelated?
4. Can ACQ-5 serve as a reliable surrogate marker for identifying patients with high T2 inflammation and SAD in settings without access to advanced testing?

By addressing these questions, our study aimed to enhance the initial treatment strategies for uncontrolled asthma in primary care, contributing to more personalized and effective asthma care.

## Methods

### Study Design and Participants

This single-center retrospective cohort study evaluated treatment-naïve adults with uncontrolled asthma who presented at our primary care clinic for initial treatment between April 2020 and March 2022. Eligible participants were adults aged ≥ 18 years who met the stringent diagnostic criteria for asthma based either characteristic respiratory symptoms (recurrent wheezing, chest tightness, dyspnea, and chronic cough) or demonstrated lung function improvement (≥ 12% and ≥ 200 mL in FEV_1_) following ICS-containing treatment.^11^ Uncontrolled asthma was defined as an ACQ-5 score of ≥ 1.5.^8^ All participants were treatment-naïve and abstained from oral corticosteroids, ICS, LABA, long-acting muscarinic antagonists (LAMA), inhaled combination therapies, antileukotrienes, theophylline, and biologics for at least six months before enrollment.

The exclusion criteria were recent pneumonia, significant cardiorespiratory comorbidities, short-acting beta-agonist (SABA) use within 6 hours before the clinic visit, pregnancy, or lactation. To minimize potential COPD and asthma-COPD overlap (ACO), we excluded patients with a smoking history ≥ 10 pack-years and symptom onset after 40 years of age.

### Data Collection and Assessment

Demographic data, ACQ-5 scores, spirometry results, and T2 biomarker levels were collected at the initial visit. Spirometry was performed using a P-370® spirometer (FUKUDA DENSHI, Tokyo, Japan) according to the Japanese Respiratory Society guidelines.^25^ Pulmonary function was assessed by measuring forced expiratory volume in 1 second (FEV_1_) and forced expiratory flow at 25% and 75% of vital capacity (FEF_25-75_) to evaluate proximal and distal airway function, respectively. All measurements were performed in triplicate according to Graham et al.,^26^ and the most technically acceptable and reproducible maneuver was selected for the analysis. Results were expressed as percentages of predicted values, with FEV_1_%pred < 80% and FEF_25-75_%pred < 65%, indicating abnormally low values.

Based on the pulmonary function test results, the participants were classified into three groups according to the following criteria.

1. Normal lung function (NLF) group:

- FEV_1_%pred ≥ 80% AND
- FEF_25-75_%pred ≥ 65%
2. Small airway dysfunction (SAD) group:

- FEV_1_%pred ≥ 80% AND
- FEF_25-75_%pred < 65%
3. Airflow Limitation (AFL) group

- FEV_1_%pred < 80% AND
- FEF_25-75_%pred < 65%

The T2 biomarkers were measured using the following equation:

1. XN-9100® automated hematology analyzer (Sysmex, Germany) for the blood eosinophil count (bEOS)
2. NIOX VERO® portable device (Circassia AB, Sweden) for fractional exhaled nitric oxide (FeNO)
3. A Phadia2500® immunoassay analyzer (Thermo Fisher Diagnostics, USA) was used for total serum immunoglobulin E (IgE)

Elevated T2 inflammation was defined as bEOS ≥ 300 cells/μL,^11^ FeNO ≥ 50 ppb,^27^ or total serum IgE ≥ 170 IU/mL.^28^

Based on previous research suggesting the benefit of high-dose ICS in patients with elevated T2 biomarkers,^21^ the participants were categorized as follows:

1. “T2-high”:

- bEOS ≥ 300 cells/μL AND
- FEF_25-75_%pred ≥ 65%
2. “T2-low” :

- bEOS < 150 cells/μL AND
- FeNO < 20 ppb
3. “T2-intermediate”: Neither “T2-high” nor ”T2-low”

### Statistical Analysis

Analyses were performed using EZR version 1.61 (Saitama Medical Center, Jichi Medical University, Saitama, Japan), a graphical interface for R (The R Foundation for Statistical Computing, Vienna, Austria).^29^ Continuous data were analyzed using the Mann-Whitney U test and are presented as medians with interquartile ranges (IQR). Categorical data were evaluated using Fisher’s exact test and are presented as frequencies and percentages. The relationships between variables were assessed using Spearman’s rank correlation coefficient. Multivariate analysis employed logistic regression with backward stepwise variable selection based on p values. Receiver operating characteristic (ROC) curves were constructed to evaluate the predictive performance, and the area under the curve (AUC) was calculated.

Differences among multiple groups were assessed using the Kruskal-Wallis test, followed by post hoc pairwise comparisons with Bonferroni correction to control for multiple testing.

Statistical significance was set at p < 0.05 (two-tailed).

### Patient and Public Involvement

This retrospective study analyzed the clinical data of patients with uncontrolled asthma who presented to a primary care clinic. Patients were not directly involved in the design, conduct, or dissemination of the research project. However, the study objectives were informed by the need to better understand and address the unmet needs of patients with uncontrolled asthma, particularly in the primary care setting where most initial treatments are provided.

Our findings highlight the potential value of assessing airway inflammation and lung function impairment to guide personalized treatment strategies in this population. Future prospective studies based on these findings would benefit from incorporating patient perspectives on intervention acceptability and outcome measures. Dissemination of results through patient communities may enhance awareness of asthma heterogeneity and the value of comprehensive assessment in treatment optimization.

## Results

### Patient Characteristics

Among the 282 patients initially evaluated for uncontrolled asthma, 192 were ultimately included in the final analysis (Table 1). Of these, 54 patients were excluded because of ongoing treatment, 10 were unable to complete spirometry or FeNO measurements, and 28 patients did not demonstrate positive bronchodilator responsiveness after > 4 weeks of ICS-containing treatment. The median age was 47 years (IQR: 36-59), and 49% of the participants were female. Thirty-two percent had childhood-onset asthma (age < 18 years), and 68% had adult-onset asthma (age ≥ 18 years). The median body mass index (BMI) was 22.9 (IQR: 21.0-25.6) kg/m^2^, and 8.3% of the patients were classified as obese (BMI ≥ 30 kg/m^2^). Most patients (88%) were categorized as requiring GINA starting treatment steps 4-5, with a median ACQ-5 score of 2.80 (IQR: 2.00-3.45).

**Table 1.**
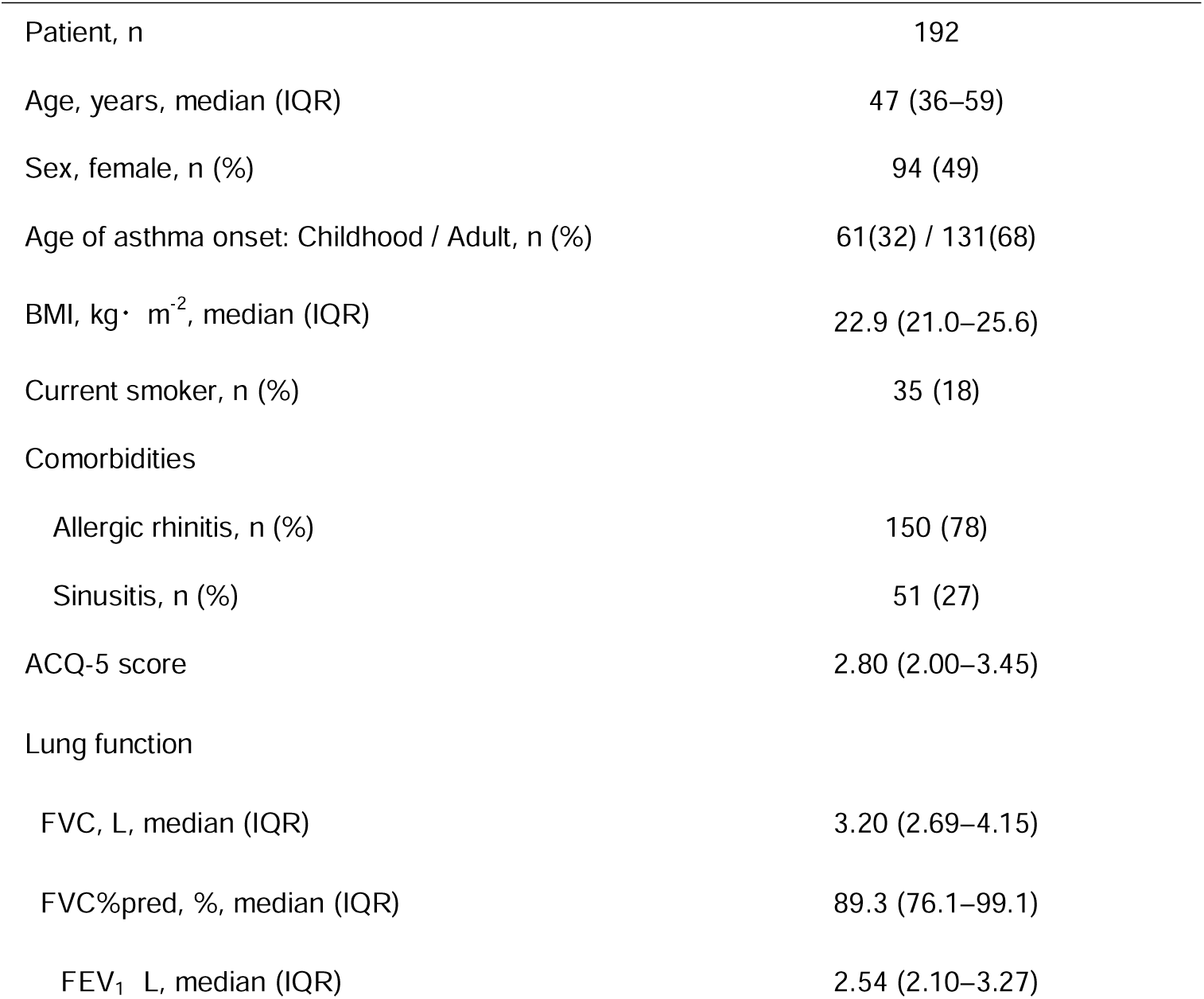
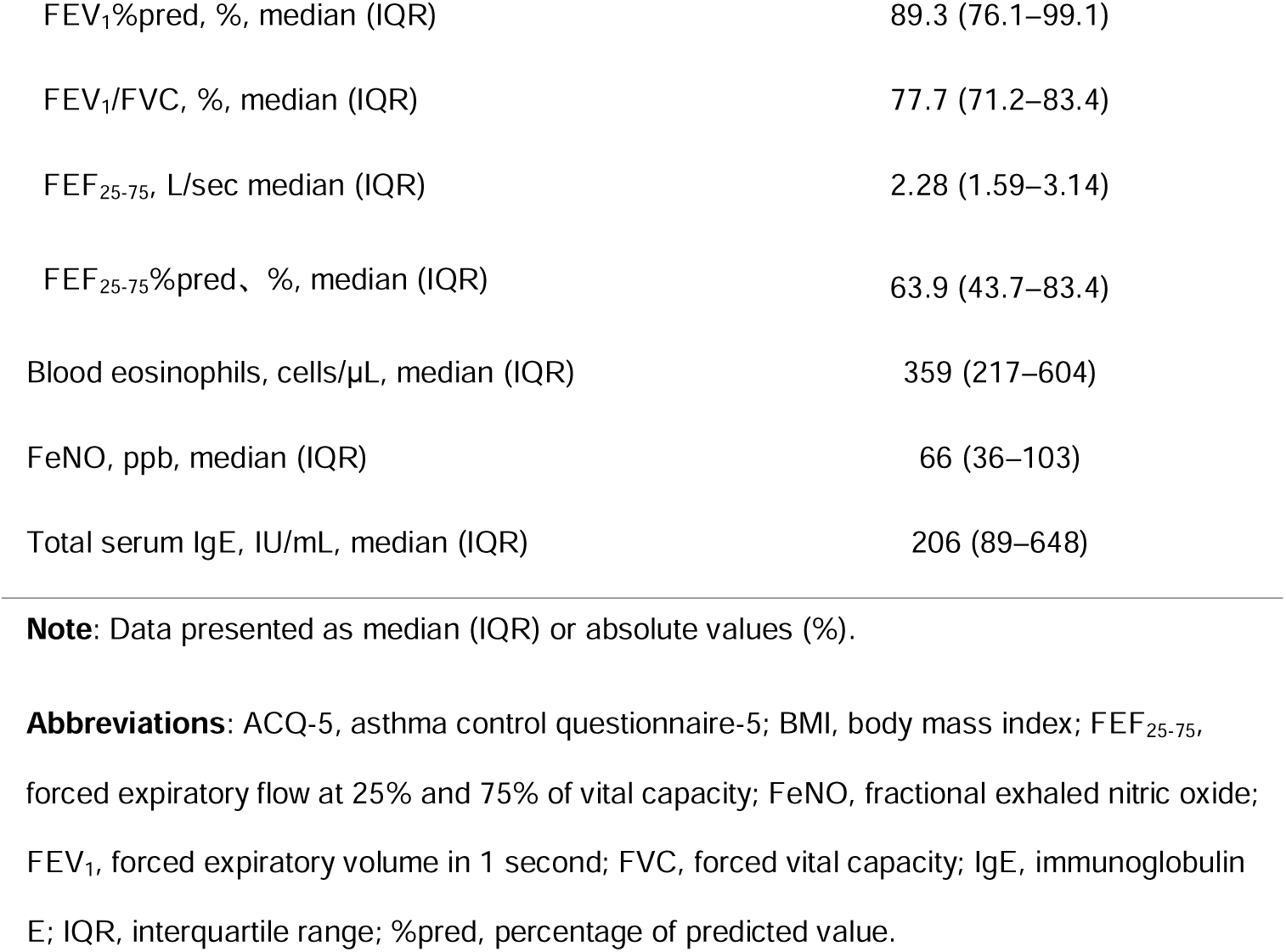
Baseline Demographics and Clinical Characteristics of Patients

### Prevalence of Elevated T2 Biomarkers

The median T2 biomarker levels were as follows: bEOS, 359 cells/μL (IQR: 217-604); FeNO, 66 ppb (IQR: 36-103); and total serum IgE, 206 IU/mL (IQR: 89-648). Figure 1 shows a substantial overlap, with 87% of patients showing at least one elevated biomarker, 63% showing two elevated biomarkers, and 29% showing all three elevated biomarkers. The phenotype distribution was: 47% “T2-high,” 51% “T2-intermediate,” and 3% “T2-low.”

**Figure 1.**
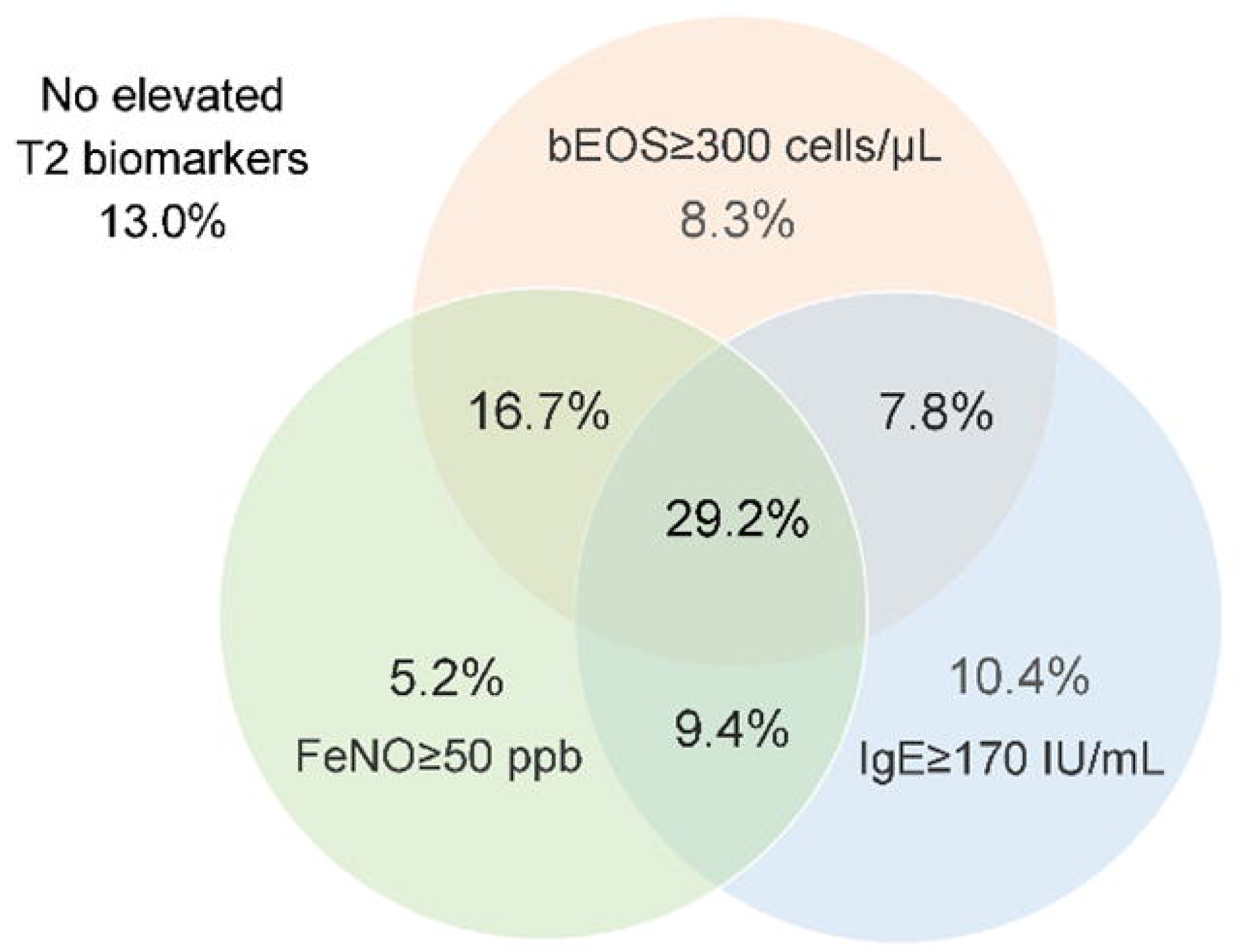
Prevalence of Elevated T2 biomarker levels. 87% of the patients had at least one elevated biomarker, 63% had two, and 29% had all three. bEOS, blood eosinophil count; FeNO, fractional exhaled nitric oxide; IgE, immunoglobulin E; T2, Type 2.

### “T2-high” Phenotype Prediction

Logistic regression analysis using age, sex, age of onset, obesity, smoking history, presence of allergic rhinitis or chronic rhinosinusitis, cough/sputum symptoms, and ACQ-5 as predictors identified an ACQ-5 score ≥ 3.0 as significantly associated with “T2-high” phenotype (odds ratio: 2.62, 95% confidence interval [CI]: 1.46-4.69, p = 0.00127). The model showed a moderate predictive ability (AUC, 0.723; 95% CI: 0.652-0.795).

### Prevalence of SAD and Association with Asthma Control and T2 Inflammation

Thirty-three percent demonstrated proximal airway obstruction (FEV_1_%pred < 80%), while 52% showed SAD (FEF_25-75_%pred < 65%). Among the patients with SAD, 40% maintained a normal FEV_1_%pred.

The median ACQ-5 scores were 2.2 (IQR: 1.6-2.8) in the “NLF group”, 3.0 (IQR: 2.2-3.4) in the “SAD group”, and 3.5 (IQR: 2.8-4.0) in the “AFL group”, demonstrating statistically significant differences among the three groups (p < 0.001). Post hoc pairwise comparisons revealed statistically significant differences between the groups (Figure 2).

**Figure 2.**
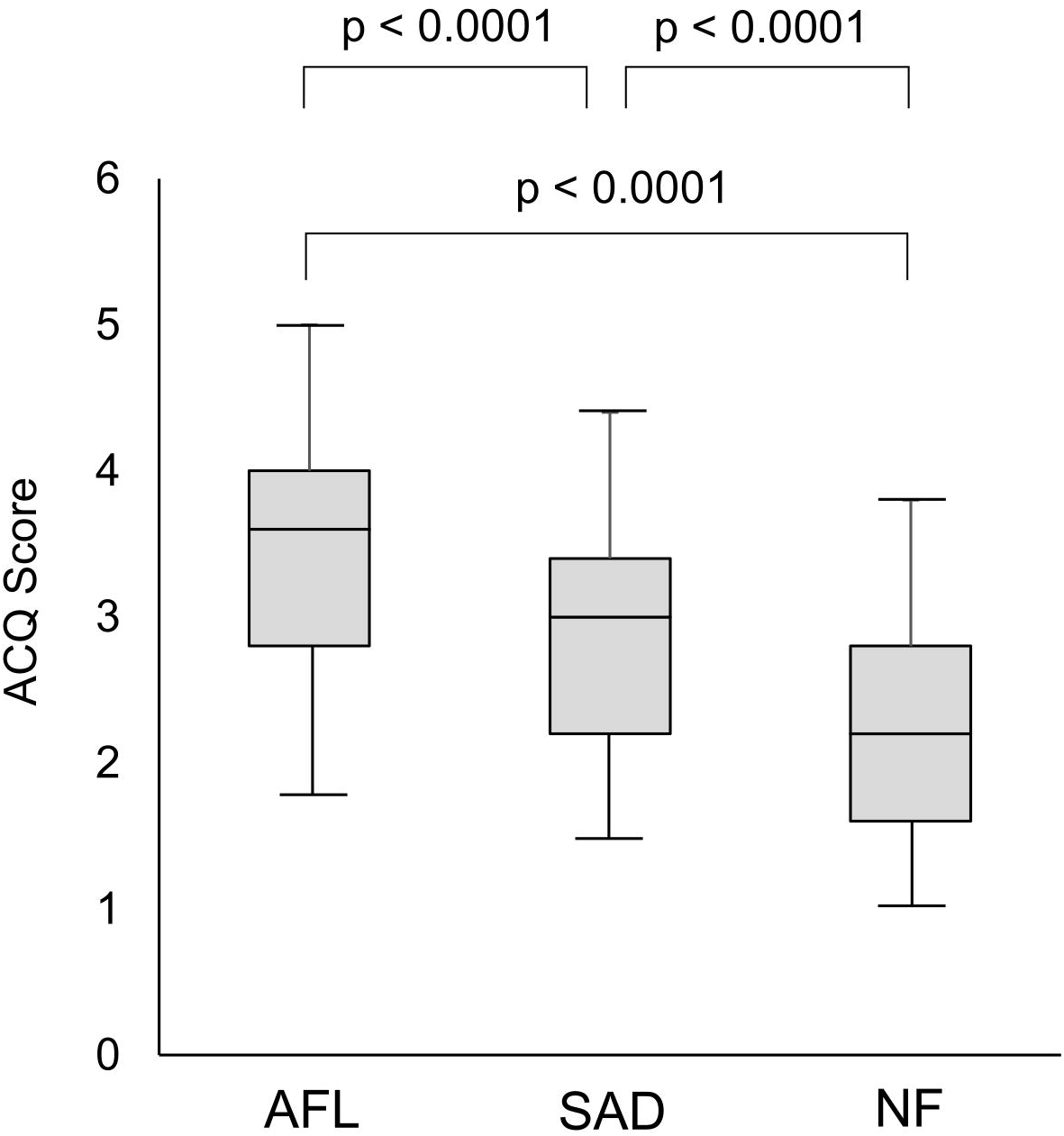
Association between Central and Peripheral Airway Dysfunction with Asthma Control. The median ACQ-5 scores (IQR) were as follows: NFL group, 2.2 (1.6-2.8); SAD group, 3.0 (2.2-3.4); AFL group, 3.5 (2.8-4.0). Statistically significant differences were observed among the three groups (p < 0.001), and post-hoc pairwise comparisons revealed significant differences between all group pairs. ACQ-5, asthma control questionnaire-5; AFL, airflow limitation; IQR, interquartile range; NFL, normal function; SAD, small airway dysfunction.

Regarding T2 biomarkers, the bEOS was significantly higher in the “AFL group” than in the “NLF group” (p = 0.033). However, there were no statistically significant differences in the FeNO or total IgE levels among the groups.

### Correlations between ACQ-5 Scores, T2 Biomarkers, and Lung Function

The ACQ-5 scores were negatively correlated with FEF_25-75_%pred (ρ = -0.583, p < 0.0001) and FEV_1_%pred (ρ = -0.567, p < 0.0001) (Table 2). Weak correlations were observed between ACQ-5 scores and bEOS (ρ = 0.231, p = 0.0014) and FeNO (ρ = 0.230, p = 0.012).

**Table 2.**
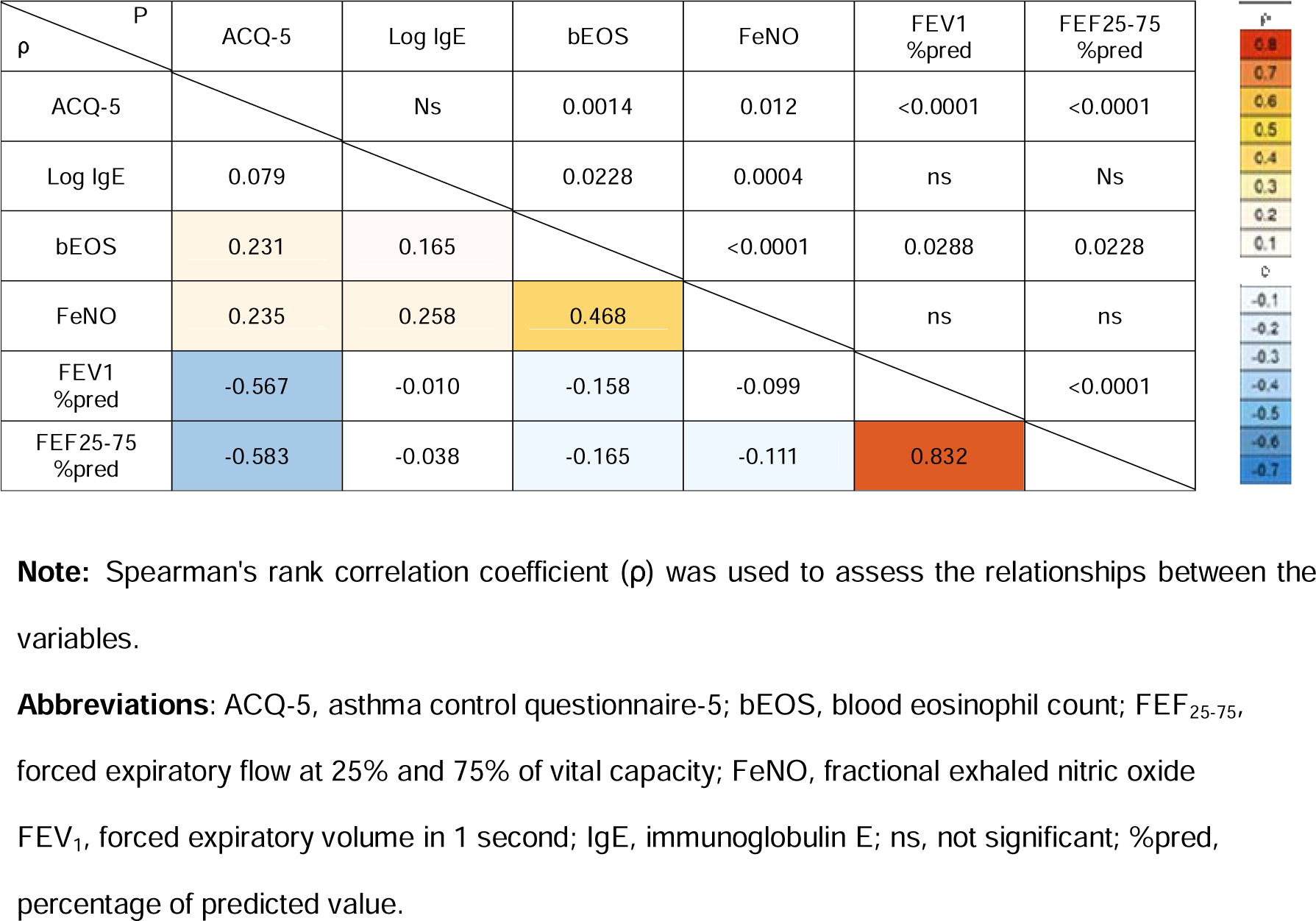
Correlations between Type 2 Biomarkers, ACQ-5 scores, and Lung Function

### Correlations between ACQ-5 Items and Lung Function

The nocturnal awakening score demonstrated the strongest negative correlation with FEF_25-75_%pred (ρ = -0.728, p < 0.0001) and FEV_1_%pred (ρ = -0.685, p < 0.0001) (Table 3).

**Table 3.**
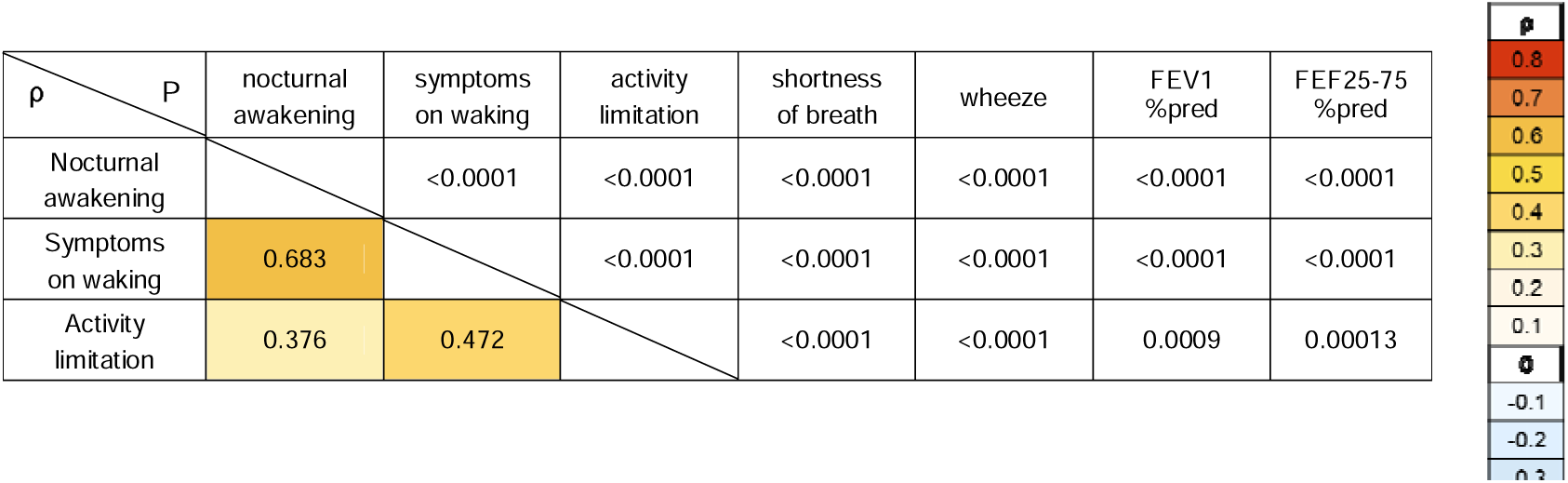
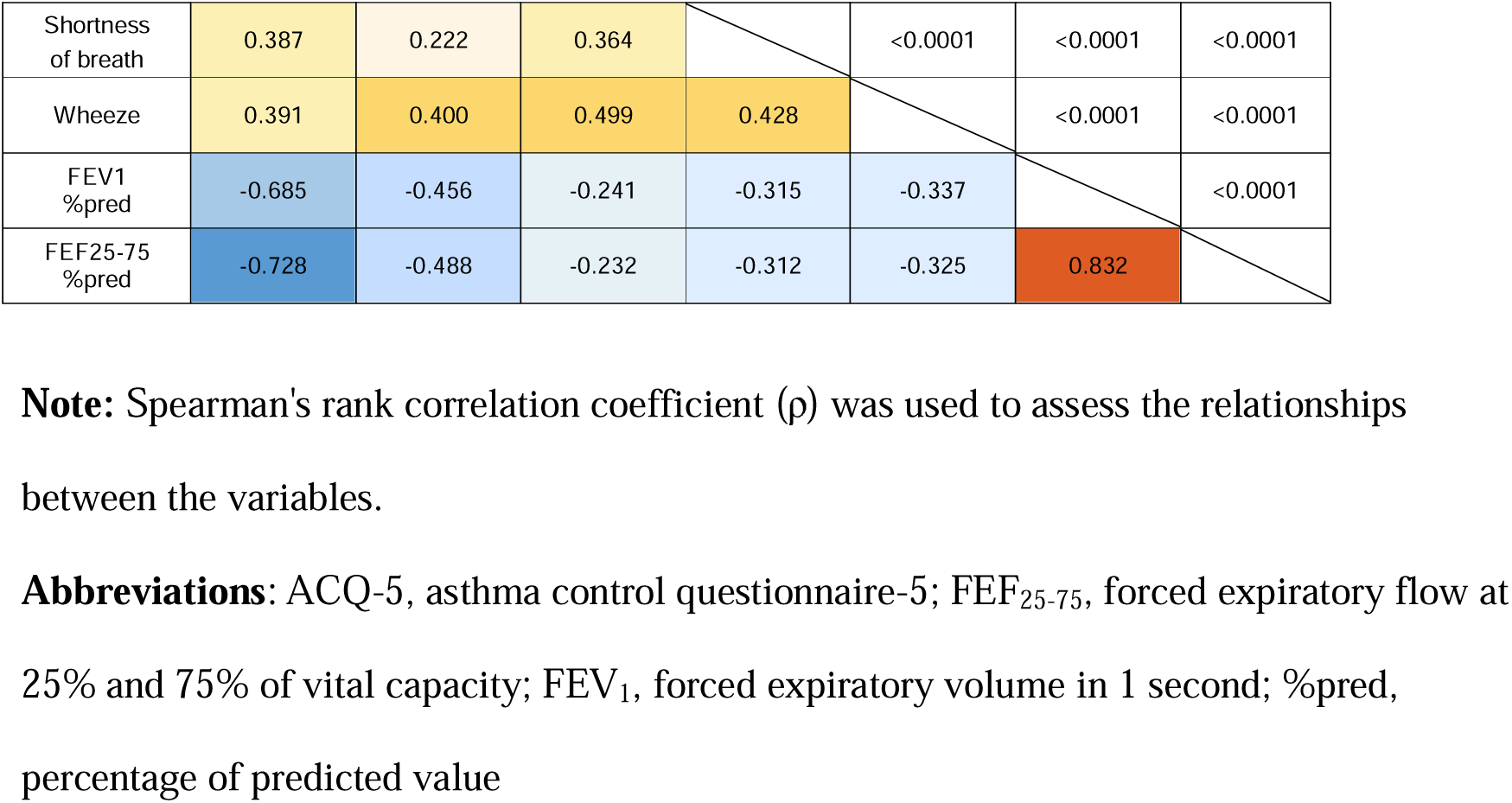
Correlations between ACQ-5 Items and Lung Function

Symptoms of waking also showed robust negative correlations with FEF_25-75_%pred (ρ = - 0.488, p < 0.0001) and FEV_1_%pred (ρ = -0.456, p < 0.0001). The other ACQ-5 items showed weak correlation (ρ < 0.4).

## Discussion

This study provides preliminary insights into the pathophysiological characteristics of uncontrolled asthma in treatment-naïve patients who receive primary care as the initial treatment. Our findings suggest a notable prevalence of elevated T2 biomarkers (87%) and SAD (52%) in this population, underscoring their contribution to uncontrolled asthma before initiating treatment. By excluding patients taking anti-inflammatory or bronchodilator medications, we ensured unbiased assessment. Furthermore, our results indicate that ACQ-5 may serve as a potential indicator of these underlying pathophysiological mechanisms, offering a practical tool for identifying T2 inflammation and SAD in primary care settings where advanced diagnostics are limited.

### Prevalence of T2 Inflammation and Implications for Treatment

A high proportion of patients (87%) exhibited elevation of at least one T2 biomarker, with 63% and 29% showing elevation of two or all three biomarkers, respectively. These findings are consistent with those of previous studies,^30^ which have demonstrated the predominance of T2 inflammation in patients with untreated uncontrolled asthma. The identification that 47% of patients met “T2-high” criteria has important therapeutic implications. A post hoc analysis of the CAPTAIN trial,^21^ suggested that high-dose ICS therapy in T2 high patients may yield superior improvements in lung function and exacerbation reduction compared with medium-dose ICS therapy. However, high-dose ICS therapy carries potential risks, including adrenal suppression^31^ and other glucocorticoid-related side effects.^32–35^ Future prospective randomized controlled trials are needed to establish the optimal initial ICS dosing strategy and treatment duration in this patient population.

### Small Airway Dysfunction and Its Clinical Significance

We identified SAD as FEF_25-75_% pred < 65% in 52% of the patients, with 40% of these individuals maintaining a normal FEV_1_%pred. This finding highlights that SAD can exist despite preserved proximal airway function and may be under-recognized in primary care settings. Our categorization of patients into the “NLF,” “SAD,” and “AFL” groups revealed progressive worsening of asthma control across these groups, with median ACQ-5 scores of 2.2, 3.0, and 3.5, respectively (p < 0.001). This stepwise increase in the ACQ-5 scores suggests that SAD represents an intermediate state between normal lung function and frank airflow limitation, potentially marking a critical window for therapeutic intervention.

The strong negative correlation between FEF_25-75_%pred and ACQ-5 scores (ρ = -0.583) demonstrates that SAD significantly impaired asthma control. Moreover, the very strong negative correlation with nocturnal awakening scores (ρ = -0.728) aligns with previous research showing marked eosinophilic infiltration in the distal airways during early morning bronchoscopy in patients with nocturnal asthma.^36^

Interestingly, although bEOS showed significant differences between the “NLF” and “AFL” groups (p = 0.033), the FeNO and total IgE levels did not differ significantly between the groups. This pattern suggests that systemic eosinophilia may parallel the progression of airway dysfunction,^37^ whereas local airway inflammation (measured by FeNO) and allergic sensitization (indicated by IgE) may be more uniformly present across the disease spectrum. This finding adds to our understanding of the relationship between T2 inflammation and airway dysfunction, indicating that different aspects of T2 inflammation may have different associations with SAD progression. Additionally, FEF_25-75_ has been shown to be the most reliable predictor of airway hyperresponsiveness to mannitol challenge ^38^ These collective findings emphasize the need for early SAD identification and targeted therapeutic approaches,^39^ particularly in patients who maintain normal FEV_1_ values but show early signs of small airway involvement.

### ACQ-5 as a Practical Screening Tool

Our findings demonstrate the value of ACQ-5 as a screening tool for both the T2-high phenotype and SAD. Logistic regression analysis revealed that an ACQ-5 score ≥ 3.0 effectively predicts T2-high status. The strong correlation between ACQ-5 scores (particularly nocturnal awakening items) and FEF_25-75_%pred suggests that elevated scores strongly indicate SAD. While current guidelines recommend PROMs for asthma control assessment, their implementation in primary care remains suboptimal.^40^

The ACQ-5 offers distinct advantages over other tools such as the Asthma Control Test (ACT),^41^ particularly in untreated patients. Its one-week recall period enables a more accurate assessment of current symptoms, and independence from SABA use makes it especially suitable for treatment-naïve patients. These features, combined with their predictive value for T2 inflammation and SAD, suggest that the ACQ-5 is an ideal tool for guiding initial treatment decisions in primary care settings.

### Limitations

This study has several limitations. First, the absence of a treatment group limits comparative analysis. However, our focus was on characterizing the pathophysiology of untreated patients, which is common in primary care settings. Second, the single-center design and relatively small cohort size may limit the generalizability and statistical power. Third, we did not directly compare the effectiveness of high-dose and medium-dose ICS in the “T2-high” subgroup. Finally, incorporating advanced diagnostics, such as forced oscillation testing (FOT),^42–44^ additional T2-related biomarkers, such as periostin,^45^ and non-T2 biomarkers, such as YKL-40,^46^ could provide a more comprehensive assessment.

## Conclusions

Our study revealed a significant prevalence of high-grade T2 airway inflammation and SAD in treatment-naïve patients with uncontrolled asthma. Notably, we identified progressive deterioration in asthma control across the spectrum from normal lung function through SAD to airflow limitation, with SAD representing a critical intermediate state. The differential patterns of T2 biomarkers across these groups, particularly the association between the blood eosinophil count and disease progression, provide new insights into the complex relationship between inflammation and airway dysfunction.

In primary care settings, where advanced assessment of airway inflammation and comprehensive lung function testing may be challenging, the integration of the ACQ-5 into routine practice offers a practical solution. This simple yet effective tool can empower clinicians to screen for and identify not only underlying inflammatory processes but also the presence of SAD, even before obvious airflow limitation develops. By leveraging ACQ-5, healthcare providers could potentially optimize asthma management strategies from the outset of patient care, with particular attention paid to early intervention in patients showing signs of SAD. This approach not only enhances the quality of care in primary settings but also paves the way for more personalized and effective asthma management. Future research should focus on validating these findings in larger multicenter studies and establishing optimal treatment algorithms based on initial ACQ-5 scores and the presence of SAD.

## Footnotes

**Contributor-ship statement:** YN, RN, TY, and AA designed the study, interpreted the data, and critically reviewed and approved the manuscript. YN developed the analytical plan for this study. All authors have read and approved the final manuscript.

## Competing interests

The authors declare no conflict of interest.

## Competing Interests

YN received honoraria from AstraZeneca K.K. and GlaxoSmithKline. RN, HY, and TA: None.

## Acknowledgments

We would like to express our sincere gratitude to the nurses at the Nakano Respiratory and Allergy Clinic for their assistance with spirometry, exhaled nitric oxide measurements, and blood collection. Their expertise and dedication were essential to the success of this study.

## Ethics approval

This study was approved by the Institutional Review Board (approval no. 22-02, dated August 30, 2022) and adhered to Good Clinical Practice guidelines and the Declaration of Helsinki. Clinical trial registration was not required because the data were retrospectively analyzed. The requirement for informed consent was waived owing to the retrospective design of the study, and the data were anonymized. However, the patients were informed about the study through clinic bulletins and websites, with the option to opt out.

## List of abbreviations

ACT: asthma control Test
ACQ-5: asthma control questionnaire-5
AFL: airflow limitation
bEOS: blood eosinophil count
AUC: area under the curve
BMI: body mass index
FEF_25-75_: forced expiratory flow at 25% and 75% of vital capacity
FeNO: fractional exhaled nitric oxide
FEV_1_: forced expiratory volume in 1 second
FOT: forced oscillation technique
FVC: forced vital capacity
ICS: inhaled corticosteroid
IgE: immunoglobulin E
IL: interleukin
IQR: interquartile range
NLF: normal lung function
ns: not significant
%pred: percentage of predicted value
PROM: patient-reported outcome measure
ROC: receiver operating characteristic
SAD: small airway dysfunction
T2: Type 2.

## Notes

### Competing Interest Statement

The authors have declared no competing interest.

### Author Declarations

This study was approved by the Ethics Committee of Nakano Respiratory and Allergy Clinic (approval no. 22-02, dated August 30, 2022). Informed consent was waived due to the retrospective nature of the study, and an opt-out system was implemented. All data were anonymized, and the study adhered to Good Clinical Practice guidelines and the Declaration of Helsinki.

